# Comparing Fourteen Consensus Biomarkers of Aging: Epigenetic Pace of Aging as the Strongest Predictor of Mortality in BASE-II

**DOI:** 10.1101/2025.11.20.25340648

**Authors:** Valentin Max Vetter, Marit Philine Junge, Christian A. Drevon, Thomas E. Gundersen, Jan Homann, Christina M. Lill, Ulman Lindenberger, Graham Pawelec, Lars Bertram, Denis Gerstorf, Ilja Demuth

## Abstract

In many countries, lifespan has been increasing faster than healthspan, leading to more years spent with late-life disease and highlighting the need for reliable biomarkers to measure biological aging and to plan personalized interventions to extend healthspan. We used data from the Berlin Aging Study II (BASE-II, 60-80 years of age at baseline, average follow-up 7.4±1.5 years, range 3.9-10.4, n=1,083) to compare 14 biomarkers of aging recently consented by an expert panel for the use as outcome measures in intervention studies: Insulin-like growth factor 1 (IGF-1), growth-differentiating factor-15 (DNA methylation derived, DNAmGDF15), high sensitivity C-reactive protein (CRP), interleukin-6 (IL 6), muscle mass, muscle strength, hand grip strength (HGS), Timed-Up-and-Go (TUG), gait speed, standing balance test, frailty index (FI), cognitive health, blood pressure, and epigenetic age (DunedinPACE). Cox proportional hazard regression analyses were performed to investigate the predictive role for all-cause mortality and to identify subgroups of the three most frequent causes of death observed in BASE-II. Results were adjusted for age, sex, lifestyle factors, and genetic ancestry. In adjusted models of all-cause mortality, HGS, IL 6, standing balance, cognitive health, and epigenetic age (DunedinPACE) significantly predicted mortality, with the epigenetic age (DunedinPACE) emerging as the strongest predictor. In contrast, CRP, Gait Speed, IGF-1, blood pressure, muscle mass, DNAmGDF15, FI and TUG were not associated with mortality. These results were corroborated in subgroup analyses stratified by cause of death. Feature selection identified a minimal biomarker set comprising muscle mass, standing balance, and epigenetic age (DunedinPACE) that predicted mortality with nearly the same discriminative accuracy (C-index = 0.63) as the full model including all biomarkers (C-index = 0.65).

## Introduction

The continuous increase in lifespan in most industrialized countries over the past few decades has led to a demographic shift towards an aging population [1]. However, this has not been sufficiently matched by an increase in healthspan, the time of life spent without disease [1–3], resulting in an estimated average 16-20% of the lifetime spent with late-life morbidity [4]. However, biological aging occurs at different rates in different individuals. Whereas some people are affected by age-related decline in physical and mental functions and have chronic diseases at an early age, others maintain good health and overall function well into old age. To assess objectively these differences in biological aging, a variety of biomarkers has been developed. One proposed application of these biomarkers is the detection of unfavorable health conditions as early as possible so that disease can be prevented in a targeted manner before they occurs rather than treated after clinical manifestation (e.g. reviewed in [5]). Moreover, biomarkers can be used to investigate the effectiveness of interventions in studies (e.g. reviewed in [6]) with shorter follow-up periods so their effects can be discerned many years before the impact on a clinical phenotype. However, due to lacking consensus on the definition of biological aging [7] and the large number of candidate biomarkers, it remains unclear which biomarkers are best suited for which purposes [5]. Thus, systematic biomarker validation was identified as a key priority [5, 8].

Accordingly, Perri and colleagues carried out a three-round Delphi procedure in an effort to reach a consensus on biomarkers of aging best suited as outcome measures for intervention studies, resulting in an “Expert Consensus Statement on Biomarkers of Aging for Use in Intervention Studies” in 2025 [9]. In the first round, experts suggested biomarkers suitable for use in intervention studies. In the following two rounds of the Delphi process, a consensus emerged among the participating experts on the following 14 biomarkers: insulin-like growth factor 1 (IGF-1), growth-differentiating factor-15 (GDF-15), high sensitivity C-reactive protein (CRP), interleukin-6 (IL 6), muscle mass, muscle strength, hand grip strength, Timed-Up-and-Go, gait speed, standing balance test, frailty index, cognitive health, blood pressure, and DNA methylation/epigenetic clocks. Most of these biomarkers have been known for a long time and/or have been studied extensively in association with health-related outcomes (e.g. ref. [10–13]) and all biomarkers have been described to be associated with mortality (see a non-exhaustive overview of respective publications in Supplementary Table 1). However, due to differences in the study populations, comparability between biomarkers can be limited and, to date, a direct comparison of these 14 biomarkers in the same cohort does not seem to exist.

As noted by Perri et al. “Expert consensus identified 14 potential biomarkers of aging, which may be used as outcome measures in intervention studies. Future aging research should identify which combination of these biomarkers has the greatest utility”[9]. To address this research gap, we first examined these 14 biomarkers for their association with mortality among Berlin Aging Study II (BASE-II) participants, a large longitudinally followed cohort, allowing direct comparison of effect sizes within the same individuals and addressing a key criterion for biomarkers of aging as defined by the American Federation for Aging Research [14]. Second, subgroup analyses stratified by cause of death are provided to inform future intervention studies in selecting appropriate endpoints for reducing cause-specific mortality. Third, we identified the minimal biomarker combination exhibiting the highest variance explanation to predict mortality in our data set. This is the first study that analyses all 14 consented biomarkers in one study sample.

## Methods

### Study population

The observational longitudinal BASE-II is an interdisciplinary study investigating healthy vs. unhealthy aging trajectories [15]. For the medical baseline assessment, participants were recruited through the Max Planck Institute for Human Development’s participant pool in Berlin, which was originally established through advertisements in local newspapers and public transportation networks. Individuals aged 60 to 80 years (older age group) and 20 to 35 years (younger age group, not included in the current analysis) were eligible for participation. Between 2009 and 2014, the medical baseline assessment (T0) was completed for 1,671 participants in the older age group [15]. On average 7.4 years later (SD: 1.5 years, range: 3.9 to 10.4 years), 1,083 older participants were followed up as part of the GendAge study [16].

All participants gave written informed consent. All assessments at baseline and follow-up were conducted in accordance with the Declaration of Helsinki and approved by the Ethics Committee of the Charité-Universitätsmedizin Berlin (approval numbers EA2/029/09, EA2/144/16, and EA2/224/21) and registered in the German Clinical Trials Registry as DRKS00009277 (BASE-II) and DRKS00016157 (GendAge). This manuscript was created in accordance with the STROBE guidelines [17].

### Biomarkers

Biomarkers investigated in this study were chosen based on the consensus statement published by Perri and colleagues in 2025 [9]. An overview of all investigated biomarkers and respective methods stratified by timepoints and use in the main as well as the sensitivity analyses are given in Supplementary Table 2. The following biomarkers were investigated:

#### IGF-1

Blood was collected on filter papers (Dried Blood Spot) and IGF-1 was measured by enzyme-linked immunosorbent assay (ELISA) by VITAS Analytical Services, Ltd. Oslo, Norway using AM-248 - Quantitation of IGF-1 in dried blood spots (DBS) using ELISA. In short, punches from DBS containing human whole blood were eluted in phosphate-buffered saline (PBS) at 4 °C overnight. After equilibration to room temperature, the eluates were analyzed using the DG100B Quantikine Human IGF-1 ELISA kit (R&D Systems, Minneapolis, USA) according to the manufacturer’s instructions.

#### CRP

Due to limited data availability, “normal” (non-high sensitivity) CRP was used in our main analyses. It was measured from serum samples using immunoturbidimetry in an accredited standard hospital laboratory (Labor Berlin - Charité Vivantes GmbH, Berlin, Germany) with the Roche/Hitachi cobas c system (COBAS integra, Roche Diagnostics, Indianapolis, USA). hsCRP, obtained by collection of blood spotted on filters (DBS) from VITAS Analytical Services, was measured by Vitas analytical services using AM-438 - Quantification of hs-CRP in DBS, Mitra sticks, whole blood and plasma using MSD Mesoscale ECL. In short, one 3.1 mm punch from dried human whole blood samples (DBS) were eluted in 60 µL kit diluent and left standing at 4°C over night. After bringing the eluate to room temperature, 10 µL of the eluate were diluted in 500 µL kit diluent and analysis was then performed on a MESO® QuickPlex SQ 120 Multiplex Imager using the V-PLEX Human CRP kit (K151STD-2) from MSD, Rockville, Maryland, USA as described in the kit manual.

#### IL6

IL6 at baseline was measured using the highly sensitive Cytometric Bead Array flex kit (BD biosciences, San Jose, US) following the manufactureŕs instruction with the addition of a further dilution of the standard measured three times to improve accuracy of the standard curves. Tracking beads and cytometer setup were monitored over time to guarantee constant performance of the flow cytometer (BD LSR-II).

IL 6 at the follow-up was measured from blood spotted on filters (DBS) by VITAS Analytical Services using AM-454 Quantification of cytokines in DBS and plasma using MSD U-PLEX platform. In short, two 3.1 mm punches from dried human whole blood samples (DBS) were eluted in 75 µL kit diluent for 2 hours at 700 rpm in room temperature. 50 µL of the eluate was used in the analysis, which was performed on a MESO® QuickPlex SQ 120 Multiplex Imager and using the custom U-PLEX Biomarker Group 1 Multiplex assay from MSD, Rockville, Maryland, USA as described in the kit manual.

#### Muscle mass

Appendicular lean mass (ALM) was measured with dual-energy X-ray absorptiometry (Hologic^®^ QDR^®^ Discovery™; Hologic, Inc., Bedford, MA, USA) by a trained technician and calculated using the system software (APEX version 3.0.1., Hologic Inc. Bedford, MA, USA). To increase comparability of results between participants, ALM was BMI-standardized. Further information on ALM in BASE-II can be found elsewhere [18].

#### HGS

Maximum HGS was measured using a Smedley Dynamometer (Scandidact, Odder, Denmark). Participants were instructed to perform a maximum isometric contraction while standing, with the elbow flexed at 90° and the shoulder in a neutral rotation. The test was performed three times with each hand and the mean of the highest value from the left and right hand were used in the analyses.

*Muscle strength* was calculated as a binary variable from continuously measured HGS using the age- and BMI-specific cut-off values defined by Fried and colleagues as part of their frailty phenotype [19].

#### TUG

Participants were instructed to rise from a standard chair, walk to a line marked three meters away, turn around, return to the chair, and sit down again. The time the participants needed to perform this task was measured in seconds using a stopwatch. Impairment in TUG was defined if the participants exceeded the pre-defined cut-off of 10 seconds [12].

*Gait speed* was measured as the time (in seconds) required to walk 4 meters on level ground. Thus, higher values reflected slower gait speed.

#### Standing balance test

The Tinetti Mobility Test [20] is a clinical assessment designed to evaluate fall risk in older individuals, comprising two parts: balance and gait. For this study, only the part evaluating the participantś balance was used involving tasks such as unsupported standing, turning, and maintaining stability when lightly pushed. Depending on the task, between 0 and 4 points were awarded and summed. A summed test value below 15 was used to define impairment.

*Frailty index* was assessed using the five criteria defined by Fried and colleagues: weight loss, exhaustion, weakness, slow walking speed, and low physical activity [19]. A point was given if the participant provided values in one of the five categories below or above a specific cut-off resulting in a point range between 0 and 5. To implement this index in BASE-II, some small adjustments are described in detail elsewhere [21]. Pre-frail and frail participants (>0 points) were considered frail for the purpose of this study which is in accordance with a procedure described in BASE-II before [21, 22].

*Cognitive health* was assessed using the Digit Symbol Substitution Test (DSST), which measures concentration, working speed, visuomotor coordination, and visual short-term memory. Participants are presented with a coding key pairing numbers with unique symbols and are asked to transcribe the corresponding symbol for each number as quickly as possible within 90 seconds. The total number of correct allocations is recorded as the test score, with higher scores indicating better cognitive performance.

*Blood pressure* was measured on both arms using an electronic sphygmomanometer (bosomedicus memory; Jung, Willingen, Germany). The mean systolic blood pressure from both arms was calculated and used for the analyses. If assessment of blood pressure on only one arm was possible/available, this value was used.

#### Epigenetic clocks

EDTA-added whole blood samples were stored at -80°C and used for DNA isolation via the “Plus XL manual kit” (LGC). DNA methylation data in samples collected at T0 (n=1,011, subgroup selected for participation at T1) and T1 (n=1,052) were obtained using the “Infinium MethylationEPIC” array, version 1 (Illumina, Inc., USA). Raw data were processed using the “Bigmelon” package in R [23]. Data processing and quality control procedures are described in detail elsewhere [24].

In our main analyses, we investigated “DunedinPACE” calculated as set out in the original publication by Belsky and colleagues [25]. As additional analyses, all main models were also calculated using Horvath’s clock [26], Hannum’s clock [27], PhenoAge [28], GrimAge [29], as well as the principal component (PC) version of these clocks [30]; and finally the 7-CpG clock [31]. The DNA methylation age acceleration (DNAmAA) of these clocks was calculated as residuals of a linear regression of DNA methylation age (DNAmA) on chronological age adjusted for leukocyte cell counts (neutrophils, monocytes, lymphocytes, and eosinophils) and used in the analyses. Further details on how these clocks were calculated in BASE-II can be found elsewhere [22, 32, 33].

#### DNAmGDF15

As directly measured GDF15 is not available in BASE-II, the DNA methylation-derived GDF15 (DNAmGDF15) was estimated from Illumina MethylationEPIC array data using the algorithm published by Lu and colleagues [29].

### Mortality

Information about the BASE-II participants’ mortality status (alive/deceased) is regularly obtained from the Berlin city registry and death certificates of deceased participants are requested from the relevant authorities. Since it is documented that the information in death certificates in Germany can be inconsistent (e.g. ref [34]) they were evaluated by a group of four BASE-II scientists, all of whom are physicians and/or biologists. Consensus agreements were reached to clarify the causality of direct, indirect and accompanying causes of death as recorded on the mortality records according to the International Statistical Classification of Diseases and Related Health Problems-10 (ICD-10) and the underlying cause of death. For the current analyses, death certificates with information about the cause of death were available for n=185 participants and in this subgroup the relationship between the top three causes of death and the 14 biomarkers were analyzed.

### Confounders

Chronological age was calculated as time in years between the participantś date of birth and date of examination at T0 and T1. Alcohol intake in g/d was assessed via a validated food frequency questionnaire [35]. Smoking behavior was assessed in one-on-one interviews as packyears. To adjust for physical activity, the first question of the Rapid Assessment of Physical Activity (RAPA [36]) questionnaire was used, which showed strong association with accelerometric activity measures available in a subgroup of BASE-II participants at T1 [37]. Genetic ancestry was quantified by the first four principal components from a principal component analyses on genome-wide SNP genotyping data.

### Imputation

All biomarkers, mortality (status and time-to-event) as well as confounders were included in the imputation procedure. Because CRP and IL 6 did not show a normal distribution, they were log_10_-transformed prior to the imputation procedure. All laboratory values underwent outlier exclusion due to suspected measurement errors. Specifically, outliers were defined as values that differed more than 4*SD from the mean. The number of excluded observations was <1% for all biomarkers. Multiple imputation were performed using R’s mice package (10 datasets, 20 iterations, method = “pmm”). A prediction matrix was produced using the *quickpred*-function with a forced inclusion of chronological age at T0 as well as sex. Mortality variables (status, time-to-event, and cause of death) were used as donors but were not imputed. Imputation results were evaluated by comparing the distribution of observed and imputed values in histograms and density plots. Algorithm convergence was checked visually. Dichotomization and definition of groups based on the application of cut-offs applied to continuously-measured variables was done for each imputed dataset individually to guarantee internal consistency within all datasets.

### Statistical Analysis

To allow easy comparison of effect sizes between biomarkers on different scales, all continuous variables were z-transformed prior to statistical analysis. This was done for each variable in each imputed dataset individually.

All statistical analyses were carried out, and all figures were drawn using the statistical computation software R, version 4.4.1. Descriptive statistics were calculated by the *CreateTableOne*-function (tableone package) using the first imputed dataset as well as the original, not-imputed dataset (Table 1, Supplementary Tables 4 - 7). Cox proportional hazard regression models were calculated using the *coxph*-function with time from biomarker measurement as underlying time scale (survival package). Regression models were adjusted for age and sex (model 0) and age, sex, alcohol consumption (g/d), smoking (packyears), physical activity (RAPA question 1), and genetic ancestry (model 1). In line with recommendations on the evaluation of biomarkers of aging [8], unadjusted regression models were calculated (model 0) and are displayed in the Supplementary Material.

**Table 1.** Descriptive statistics of participants from BASE-II from the first imputed dataset. Due to limited availability at T0, values measured at T1 are shown for methylation derived variables (epigenetic clock, DNAmGDF15) and IGF-1. Original observations are shown in Supplementary Table 5 and 7.

| Variables | Level | n | Mean/n (%) | SD | Min | Max |
| --- | --- | --- | --- | --- | --- | --- |
| Chron. Age (T0) |  | 1671 | 68.8 | 3.7 | 60.2 | 84.6 |
| Chron. Age (T1) |  | 1083 | 75.6 | 3.8 | 64.9 | 94.1 |
| Sex | men |  | 809 (48.4) |  |  |  |
|  | women |  | 862 (51.6) |  |  |  |
| CRP (log, T0) |  | 1671 | 0.1 | 1.1 | -1.9 | 3.9 |
| IL 6 (log, T0) |  | 1671 | 1.2 | 0.5 | 0 | 3.4 |
| Muscle Mass (BMI-standardized ALM, T0) |  | 1671 | 0.8 | 0.2 | 0.4 | 1.5 |
| Hand Grip Strength (HGS, T0) |  | 1671 | 34.0 | 9.7 | 9.5 | 64.5 |
| Gait Speed (T0) |  | 1671 | 3.6 | 0.8 | 2.1 | 9.6 |
| Cognitive Health (DSST, T0) |  | 1671 | 44.4 | 8.5 | 5 | 74 |
| Blood pressure (systolic, T0) |  | 1671 | 143.7 | 18.8 | 78 | 237 |
| Insulin-Like Growth Factor (IGF-1, T1) |  | 1083 | 110.4 | 35.6 | 32 | 243 |
| Epigenetic Clock (DunedinPACE, T1) |  | 1083 | 1.1 | 0.2 | 0.7 | 1.6 |
| GDF15 (DNAmGDF15, T1) |  | 1083 | 1158.4 | 123.5 | 837.8 | 1613.8 |
| Muscle Strength (T0) | Not impaired |  | 1578 (94.4) |  |  |  |
|  | Impaired |  | 93 (5.6) |  |  |  |
| Timed-Up-and-Go (TUG, T0) | Not impaired |  | 1479 (88.5) |  |  |  |
|  | impaired |  | 192 (11.5) |  |  |  |
| Standing Balance Test (Tinetti Test part1, T0) | No impaired balance |  | 1462 (87.5) |  |  |  |
|  | Possibly impaired balance |  | 209 (12.5) |  |  |  |
| Frailty Index (Fried Frailty Index), T0) | Not frail |  | 1141 (68.3) |  |  |  |
|  | Frail |  | 530 (31.7) |  |  |  |
| Alcohol (T0) |  | 1671 | 14.9 | 18.5 | 0.4 | 166.6 |
| Smoking (packyears, T0) |  | 1671 | 10.4 | 17.6 | 0 | 148 |
| Physical Activity (RAPA, T0) | no |  | 1518 (90.8) |  |  |  |
|  | yes |  | 153 (9.2) |  |  |  |

The *with*-and *pool*-function (mice package) were used to calculate the models individually in each of the imputed datasets and subsequently pool the results employing Rubin’s rules [38]. The proportional hazard assumption was tested in the first imputed dataset using the *cox.zph*-function and by visually inspecting the Schoenefeld residuals plotted over time using survminer’s *ggcoxzph*-function. The pooled C-index across all imputed datasets was calculated using the *cindex*-function (miceafter package). The minimal biomarker model with the best performance in predicting mortality in this dataset was selected using the *stepwise*-function form the StepReg package (strategy = “subset”, metric = “AICc”, sle = 0.15). Because methylation data at baseline was only available for the participants who participated also in the follow-up assessment, the regression models including all available markers at baseline were restricted to this subgroup (n=1082). To compare the value added by the biomarkers with respect to discriminate mortality to a basic clinical model, Cox regression models, including only the confounders of model 1 and 3 (no biomarker of aging), were calculated as well. Results of the regression analyses were plotted as forest plots using R’s ggplot package. To illustrate differences in the survival probability between groups stratified by the investigated biomarkers, Kaplan-Meier curves were calculated from the first imputed dataset using the *ggsurvplot*-function (survminer package).

## Results

### Study population

A total of 1,671 participants aged 60 years and older were examined during the baseline medical assessment (T0, between 2009 and 2014, mean age = 68.8 years, SD = 3.7 years, 51.6% women). On average 7.4 years later, 1,083 participants were followed up as part of the GendAge study (T1, mean age = 75.6 years, SD = 3.8 years). As of October 2025, a total of 316 deaths had been recorded. Of these, 126 occurred between the baseline and T1 examinations, and 190 occurred after T1. Among those who died after T1, 90 had participated in both the baseline and T1 examinations (Figure 1). Descriptive statistics of the investigated biomarkers are displayed in Table 1 and Supplementary Tables 4-7. Correlation was generally low between biomarkers (r<|0.4|, Supplementary Figure 1). As of October 2025, the cause of death is known from death certificates in a subset of 185 participants. The most frequent causes of death were neoplasms (ICD-10: C00-D48, n=78, 43.3%) followed by diseases of the circulatory system (ICD-10: I00-I99, n=46, 25.6%) and diseases of the nervous system (ICD-10: G00-H95, n=18, 10%). All other individual causes of death occurred in 5% of all cases or less and were not investigated separately in this study (Supplementary Table 3).

**Figure 1.**
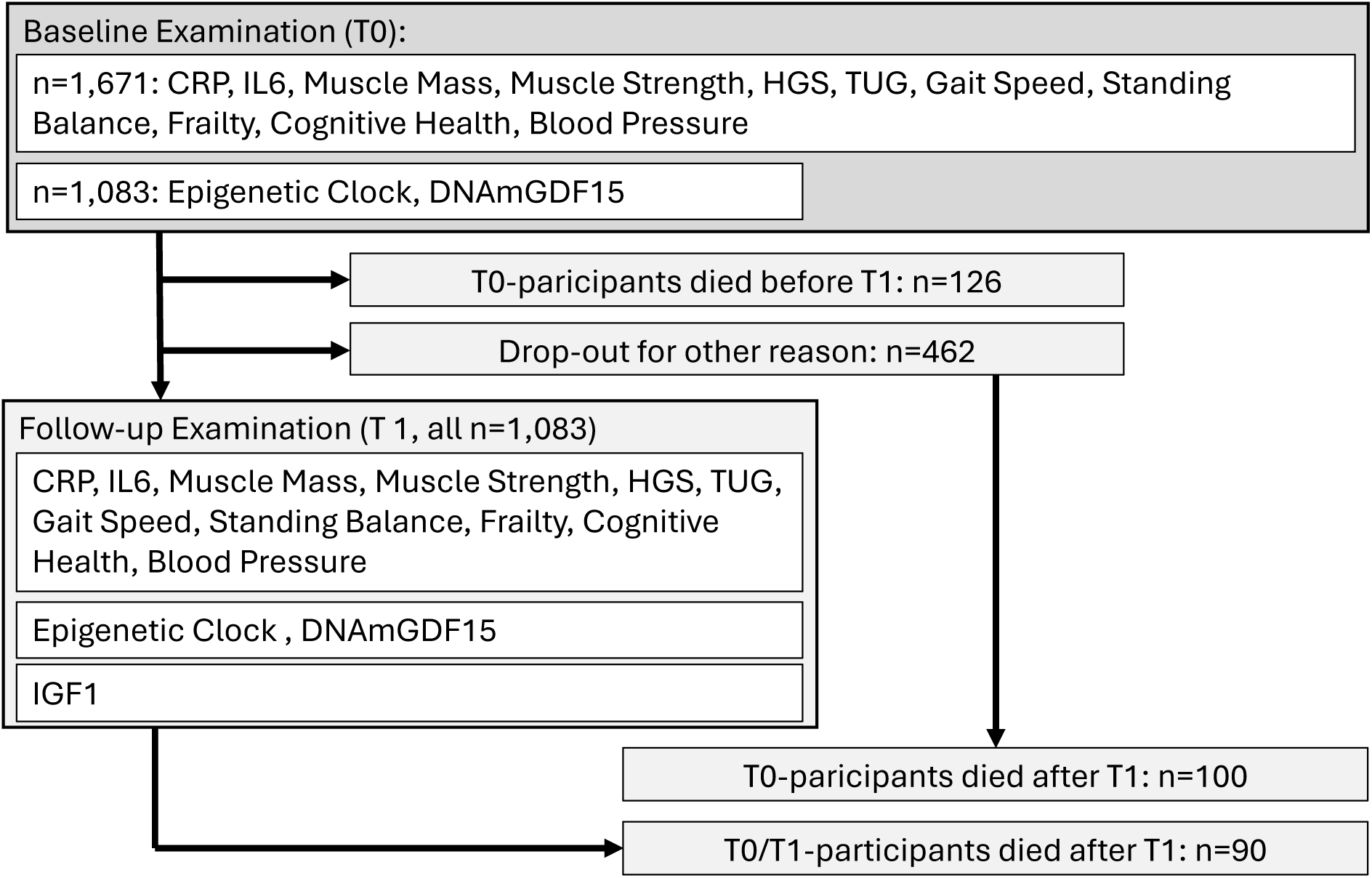
Flow chart of the availability and sample size for biomarkers at baseline (T0) and follow-up (T1) examination investigated. DNA methylation data at T0 was only available for participants who were also part of T1. IGF-1 was only available at T1.

### Association between mortality and individual biomarkers of aging

To investigate associations between the 14 biomarkers identified in the consensus procedure published by Perri and colleagues [9] and mortality, Cox proportional hazard regression models were calculated. A statistically significant association with mortality was found in the age- and sex-adjusted regression models (model 1) for all biomarkers except IGF1, Gait Speed, Blood Pressure, DNAmGDF15, Muscle Mass, and TUG. After additionally adjusting for alcohol consumption, smoking (packyears), physical activity and genetic ancestry (model 2) the epigenetic clock, IL6, cognitive health, HGS, Muscle Strength and Standing Balance remained statistically significantly associated with mortality (Figure 2A, Supplementary Table 8). Kaplan-Meier curves of the biomarkers with the strongest association to mortality are displayed in Figure 3. In sex-stratified subgroup analyses, generally similar effect sizes compared to the whole group were observed with stronger effects in the subgroup of men (Supplementary Table 8). For example, whereas no association was found between HGS and mortality in women, we observed a fairly strong association with mortality in men (HR:0.68, 95%CI: 0.54-0.84). This sex difference was statistically significant (p-value for Biomarker-Sex-Interaction: p ≤0.049, Supplementary Table 8). We repeated all regression models of biomarkers at T0 with available values measured at T1 as sensitivity analyses to enable a direct comparison for variables which were only assessed at T1 (Supplementary Table 9).

**Figure 2.**
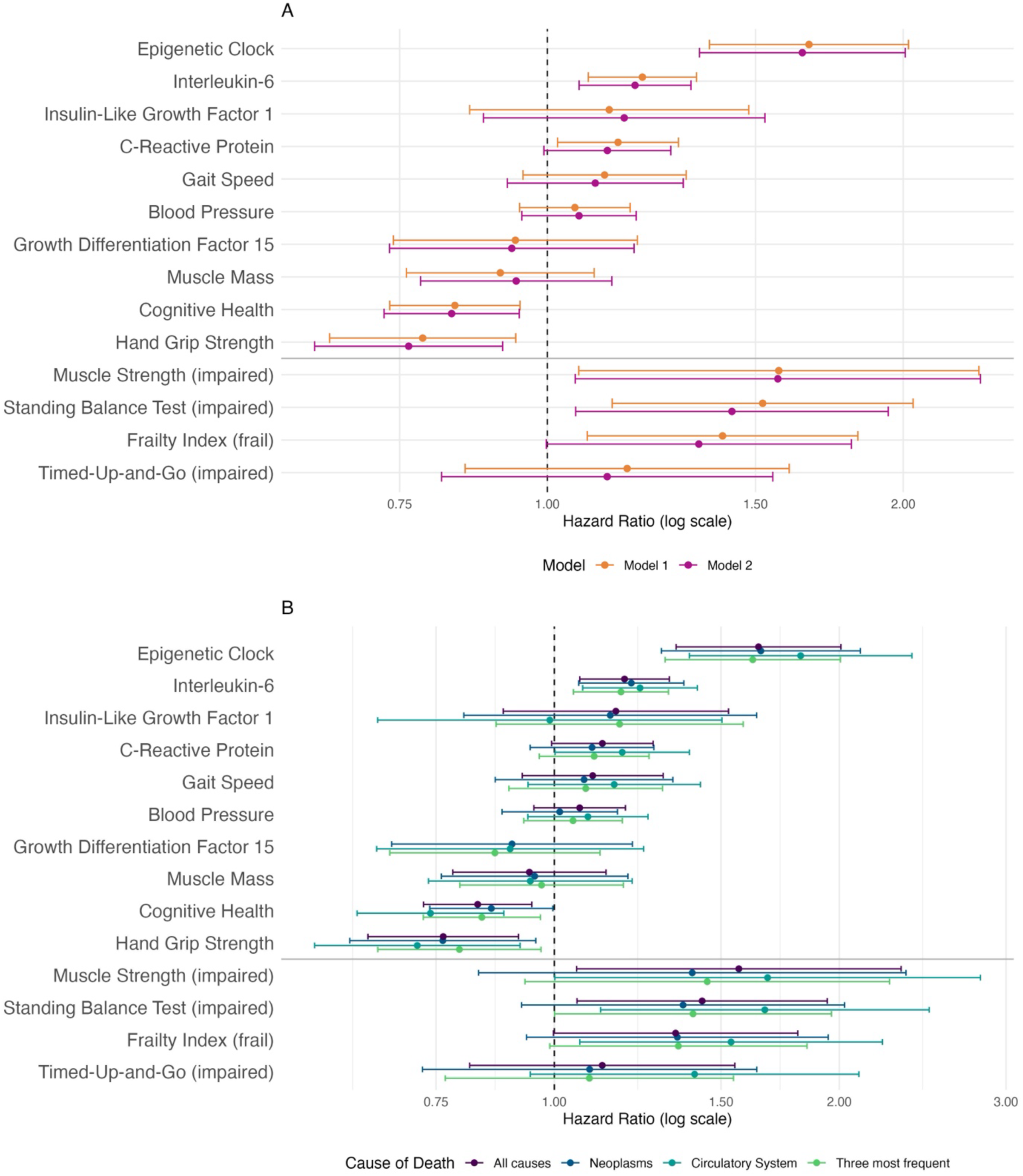
A: Forest plot of results from Cox proportional hazard regression analysis of mortality according to biomarkers of aging. All biomarkers were measured at T0 (n=1,671 participants, n=316 deaths) except for DNA methylation-derived variables (epigenetic clock (DunedinPACE) and DNAmGDF15) and IGF-1, which were measured at T1 (n=1,083 participants, n=90 deaths) on average 7.4-years later. All continuously scaled variables were normalized to allow direct comparison of effect sizes across different scales. Model 0: unadjusted; Model 1: age, sex; Model 2: Model 1 + alcohol, smoking, physical activity, genetic heredity (PC1-PC4). B: Forest plot of results from a fully-adjusted Cox proportional regression analysis (model 2) of mortality according to biomarkers of aging in subgroups defined by cause of death as obtained from the death certificates in a subgroup of BASE-II participants (n=185). All biomarkers were measured at T0 except for DNA methylation-derived variables (epigenetic clock (DunedinPACE), DNAmGDF15) and IGF-1 which were measured at T1. The values of the all-cause group are shown for easy comparison with subgroup results and are identical to values shown in A (model 2).

**Figure 3.**
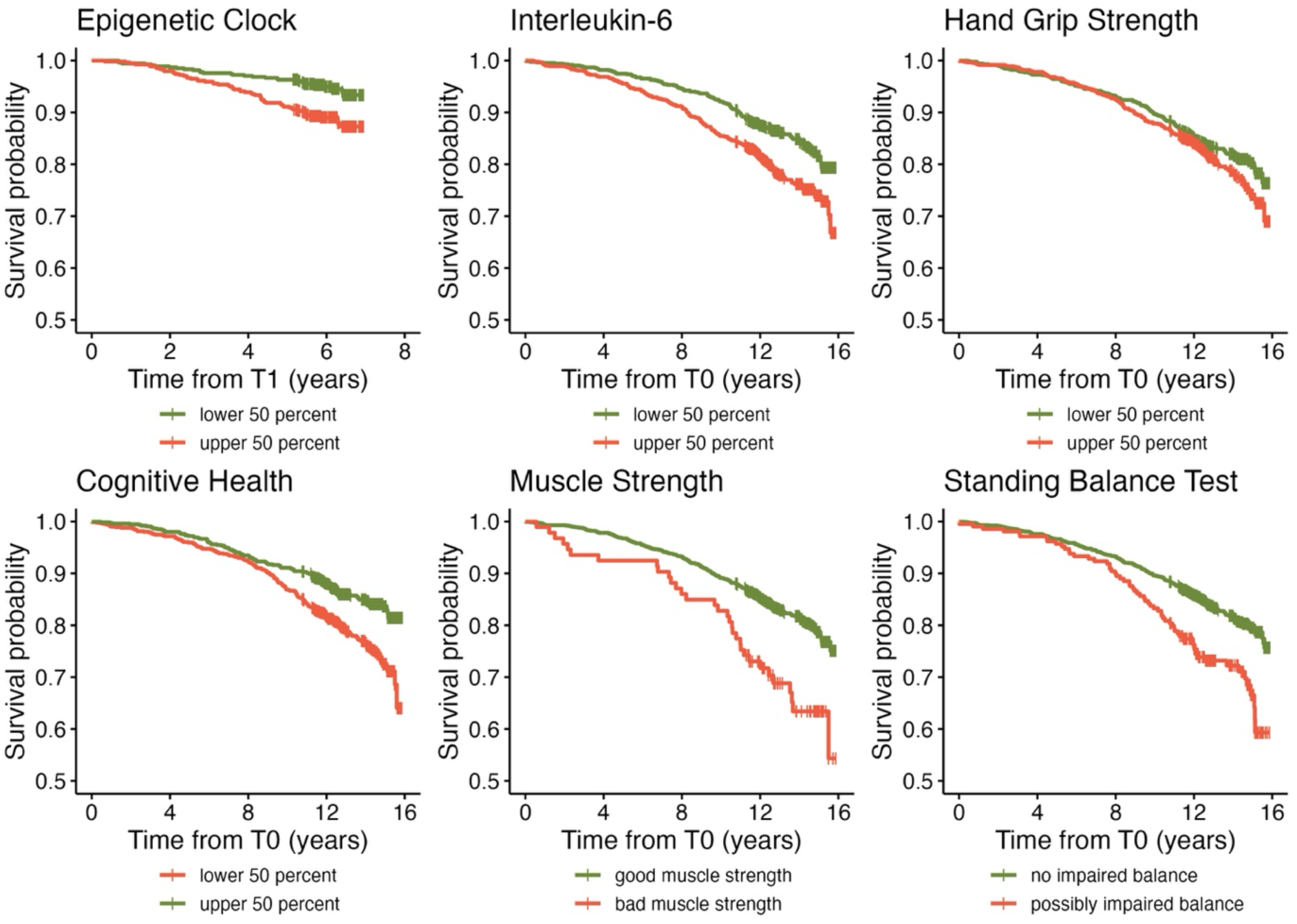
Kaplan-Meier analysis of biomarkers with the strongest association with mortality. Due to an overall low mortality rate in the sample, the y-axis limits were adjusted to improve readability of the figure. All biomarkers were assessed at T0 (n=1,671 participants, n=316 recorded deaths) except for the epigenetic clock which was derived from sample collected as part of T1 (n=1,083 participants, n=90 recorded deaths).

### Analyses stratified by cause of death

To investigate cause of death as a possible effect measure modifier, subgroup analyses were conducted on participants who died from diseases of the circulatory system, neoplasms, and the three most frequent causes of death (diseases of the circulatory system, neoplasms, and diseases of the nervous system; Supplementary Table 3). The epigenetic clock (DunedinPACE), IL 6, cognitive health (DSST), and HGS, remained statistically significantly associated with mortality among all subgroups (Figure 2B, Supplementary Tables 10, 11). Across all these biomarkers, the strongest associations were found in the subgroup of participants who died from diseases of the circulatory system. We repeated all regression models of biomarkers at T0 with values measured at T1 as sensitivity analyses to allow a direct comparison of effect sizes with biomarkers which are only available at T1 (Supplementary Table 11).

### All biomarkers of aging and best minimal set of biomarkers to predict mortality

Next, a Cox regression model including all available biomarkers of aging at T0 (Table 2, “Full Model”) was calculated to evaluate the joint ability of all biomarkers (except IGF-1, which was not measured at T0) to predict mortality. Analyses were limited by the availability of baseline DNA-methylation data, which was present for the subgroup of 1,083 participants who were also part of the follow-up examination (n=90 recorded deaths) The full Cox regression model had a C-index of 0.65. Including the confounder variables of model 1 (C-index: 0.704) and model 2 (C-index: 0.720) increased the discrimination of the Cox regression model further by up to 6.9 percentage points. Adding all available biomarkers of aging to a Cox regression model including only the confounding variables of model 1 and 3 increased the C-index by 3.9 and 4.0 percentage points (Supplementary Table 12).

**Table 2.** Cox proportional hazard regression with all (“Full Model”) or a minimal subset (“Minimal Model”) of markers. All markers in this analysis were assessed at baseline. Only participants who were also part of T1 are included in this analysis (n=1,083, n=90 deaths). Left truncation of the data was included in the regression analysis. As no IGF-1 values are available at T0, this biomarker was not part of this model.

| **Biomarker** | **HR** | **lower 95% CI** | **higher 95% CI** |
| --- | --- | --- | --- |
| **Full Model** |  |  |  |
| Epigenetic Clock | 1.39 | 1.11 | 1.74 |
| DNAmGDF15 | 1.01 | 0.81 | 1.25 |
| CRP | 0.96 | 0.75 | 1.23 |
| IL 6 | 1.11 | 0.89 | 1.39 |
| Muscle Mass | 1.26 | 0.93 | 1.71 |
| Hand Grip Strength | 0.93 | 0.68 | 1.27 |
| Gait Speed | 1.04 | 0.76 | 1.42 |
| Cognitive Health | 0.92 | 0.73 | 1.17 |
| Blood Pressure | 1.02 | 0.83 | 1.26 |
| Muscle Strength (impaired) | 1.28 | 0.49 | 3.37 |
| Timed-Up-and-Go (impaired) | 1.40 | 0.66 | 2.98 |
| Standing Balance Test (impaired) | 1.57 | 0.85 | 2.91 |
| Frailty Index (frail) | 0.96 | 0.50 | 1.84 |
| **Minimal Model** |  |  |  |
| Epigenetic Clock | 1.45 | 1.19 | 1.77 |
| Muscle Mass | 1.21 | 0.98 | 1.49 |
| Standing Balance Test (impaired) | 1.72 | 0.98 | 3.01 |

To determine the best subset of biomarkers for predicting mortality in BASE-II, a feature-selection approach was applied, considering all possible combinations of variables in the Cox regression models. The best-fitting model included the epigenetic clock (DunedinPACE), muscle mass, and performance on the Standing Balance Test (Tinetti Test, Part 1). A Cox regression model with these biomarkers provided only slightly decreased discriminatory power (C-index 0.63) compared to the full model (C-index: 0.65) with only a fraction of the biomarkers. Adding this minimal set of variables to the Cox regression models including variables in model 1 and 3, increased their discrimination both by 2.5 percentage points (Supplementary Table 12).

## Discussion

In this study, we used data obtained from 1,671 participants of the BASE-II to compare 14 biomarkers of aging which were selected through a comprehensive consensus procedure and previously published by Perri and colleagues [9] as outcome variables for anti-aging intervention studies. This is the first study that investigates and compares all 14 biomarkers in the same, well-described sample of participants. The analyses revealed individual strengths and weaknesses of these biomarkers concerning mortality. Epigenetic clock (DunedinPACE), IL6, cognitive health, HGS, Muscle Strength, and Standing Balance test were statistically significantly associated with mortality with effect sizes generally in the same range as reported in the literature (Supplementary Table 2). The epigenetic clock (DunedinPACE) showed the strongest association, and this was also observed in subgroup analyses stratified by cause of death. On the other hand, after adjusting for all confounding variables, CRP, Gait Speed, IGF-1, blood pressure, muscle mass, DNAmGDF1, Frailty and TUG were not associated with mortality. These results were observed for both all-cause mortality and cause-specific death. However, the lack of a (strong) association with mortality does not necessarily reduce the value of these variables as biomarkers of biological aging. Due to the increasing time with disease in later life, markers that are less predictive of mortality but can discriminate between those who age healthily and those who do not still hold value. Also, the instruments/methods used in this study to assess the biomarkers suggested by Perri and colleagues are often only one of numerous possible options to derive the respective variables. For example, although we used the DSST to assess *cognitive health*, this construct can be operationalized using numerous other cognitive measures and tests. Further studies are needed evaluating these 14 biomarkers of aging in the same participants not only in the context of mortality but also other age-associated outcomes, to gain a better understanding of their individual strengths and limitations.

In a final step, we investigated the joint ability of constellations of the pre-selected biomarkers of aging to predict mortality. Overall, the correlation among biomarkers was weak (r≤0.39, Supplementary Figure 1). Comparing a model containing all biomarkers of aging available at T0 to a model containing a minimal subset (epigenetic clock (DunedinPACE), Muscle Mass, and Standing Balance Test) the full cluster of markers seemed to provide only limited additional value of 3.5 and 2.5 percentage points, respectively, as compared to a basic model including age and sex. This implies that despite only weak inter-correlations, the markers seem to convey largely overlapping prognostic information. Moreover, the selection of variables in the minimal model is by no means final. Using a different selection procedure or a different study sample might identify different sets of optimal variables. In this context, we note that the high predictive value of the Standing Balance Test corroborates earlier analyses of the Berlin Aging Study (the predecessor study of BASE-II). [39]

Whereas the consensus on the 14 markers of aging was an important and highly valuable contribution to the field, the identified markers are only qualitatively described in the publication by Perri and colleagues. Thus, for most variables, no specific standardized instruments are defined. Additional studies using different methods to assess the proposed biomarkers of aging are needed, thereby providing information on which instruments are best suited as outcome measures in anti-aging intervention studies.

Strengths of this study include the availability of all 14 biomarkers of aging in the same large sample of older participants. Follow-up of up to 16 years provides a sufficient period to assess mortality for variables assessed during the BASE-II baseline assessment (T0). High quality of the cause of mortality data was ensured by an internal review procedure including an interdisciplinary team of scientist. By using the multiple imputation approach, all variables are compared within the same group of participants, which increases the comparability of effect sizes between variables. Nonetheless, this study has several limitations. First, data availability with respect to some biomarkers (DNAm derived variables, IGF-1) at baseline was limited. Thus, the follow-up time as well as the mean age differs between regression models investigating these biomarkers and all others. Second, for technical reasons, the cause of death was only available for a subgroup of participants and future studies with fully available information on cause of death are needed to explore its role for associations between biomarkers of aging and death. Third, regression models used to investigate the joint discriminative power of the available biomarkers of aging were only calculable in a subgroup of BASE-II participants. The small sample size reduced the statistical power of this study. Fourth, no adjustments for multiple testing were made.

## Conclusions

Among the 14 consensus biomarkers suggested by Perri and colleagues and here directly compared in one study sample for the first time, hand grip strength, IL 6, standing balance, cognitive health, and epigenetic age (DunedinPACE), were associated with mortality. Epigenetic age (DunedinPACE) showed the strongest and most consistent association with mortality. A minimal joint biomarker model including muscle mass, standing balance and epigenetic age (DunedinPACE) showed similar accuracy (C-index = 0.63) as the full model including all biomarkers (C-index = 0.65) suggesting the presence of a substantial degree of overlap between the investigated biomarkers with respect to mortality prediction. Further studies validating the suggested markers within additional single cohorts are needed to explore further and confirm biomarkers of human aging, particularly in diverse populations [40].

## Data availability

Due to concerns for participant privacy, data are available only upon reasonable request. Please contact Ludmila Müller, scientific coordinator, at, for additional information.

## Ethics

All participants gave written informed consent. The Ethics Committee of the Charité – Universitätsmedizin Berlin approved the study (approval numbers EA2/029/09 and EA2/144/16). The study was conducted in accordance with the Declaration of Helsinki and was registered in the German Clinical Trials Registry as DRKS00009277.

## Author contributions

Conceptualization: V.M.V, I.D.; Data curation: V.M.V., I.D.; Formal analysis: V.M.V; Investigation: V.M.V, I.D.; Methodology: V.M.V, U.L., D.G., I.D.; IGF1, hsCRP, and IL6 analyses: T.E.G.; Project administration: V.M.V, I.D.; Resources: D.G., L.B., I.D.; Supervision: D.G., I.D.; Visualization: V.M.V; Writing - original draft: V.M.V, I.D.; and Writing - review & editing: all authors.

