## Supplementary Table 1-2 and Supplementary Figure 1 for "Comparing Fourteen Consensus Biomarkers of Aging: Epigenetic Pace of Aging as the Strongest Predictor of Mortality in BASE-II"

Supplementary Table 1: Non-exhaustive overview of selected publications reporting on the association between the investigated biomarkers of aging and mortality.

| Marker | Publication | Sample Size<br>Number of Deaths | Instrument | Results |
|---|---|---|---|---|
| insulin-like growth factor 1 | (De Giorgi et al. 2022) | N=337 (19.3% women)<br>N=41<br>patients with moderately or severely HFrEF, enrolled in the T.O.S.CA. registry | IGF-1 | Cox Regression: HR: 0.42, 95%CI:0.23-0.77 |
| | (Chen et al. 2023) | N=685 (28% women)<br>N=208<br>Patients with Chronic Kidney Disease | IGF-1 | Competing risk regression:<br>high HGS + Low IGF-1: ref. group (SHR: 1)<br>high HGS + High IGF-1: SHR: 1.1, 95%CI: 0.66-1.83<br>low HGS + High IGF-1: SHR: 2.21, 95%CI:1.37-3.55<br>low HGS + low IGF-1: 2.76, 95%CI: 1.79-4.27 |
| | (Rahmani et al. 2022) | Meta-Analysis of 19 studies<br>N=30,876 | IGF-1 | random effects model with a restricted maximum likelihood heterogeneity variance estimator: high vs. low IGF-1: HR:0.84, 95%CI: 0.68–1.05<br>U-shaped relationship:<br>low vs. middle: IGF-1: HR: 1.33, 95%CI: 1.14–1.57)<br>high vs. middle IGF-1: HR:1.23, 95%CI: 1.06–1.44 |
| growth-differentiating factor-15 | (Daniels et al. 2011) | N=1391 (61% women)<br>N=436<br>Rancho Bernardo Study participants | GDF-15 | Cox proportional hazard regression: HR per SD $log_{10}$ units: 1.5, 95%CI:1.3-1.8) |
| | | N=14577<br>N= 1067<br>Stabilization of Atherosclerotic Plaque by Initiation of | GDF-15 | Cox proportional hazard regression: Highest vs. lowest quartile of GDF-15:HR 1.85, 95%CI: 1.53-5.41 |

| | | | | |
|---|---|---|---|---|
| | | Darapladib Therapy Trial (STABILITY) | | |
| high sensitivity C-reactive protein | (Li et al. 2017) | Meta-Analysis<br>N=83,995 | hsCRP | Highest vs. lowest category: RR:1.75, 95%CI: 1.55–1.98 |
| | (Boekholdt et al. 2006) | N=3272<br>N=987<br>The EPIC-Norfolk prospective population study 1993–200 | CRP | Confounder adjusted logistic regression: highest vs. lowest CRP-Quartile OR: 2.92, 95%CI: 1.83–4.67 |
| interleukin-6 | (Lee et al. 2012) | N=1843<br>N=978<br>The Rancho Bernardo Study | IL6 | Cox proportional hazard regression, confounder adjusted: HR:1.48, 95%CI: 1.33–1.64 |
| | (Puzianowska-Kuźnicka et al. 2016) | N=3750 (47.8% women)<br>1-year mortality rate: 6.6%<br>PolSenior study | IL6 | Cox proportional hazard regression, unadjusted: HR: 1.077 per each pg/mL, 95%CI:1.068–1.086 |
| muscle mass | (de Santana et al. 2021) | Meta-Analysis<br>N=10028 | appendicular skeletal muscle mass index (ASMI) | SMD between dead and living participants: ASMI<br>SMD = −0.18, 95%CI: −0.23 to −0.12 |
| | (Abramowitz et al. 2018) | N=11687<br>N=1819<br>National Health and Nutrition Examination Survey 1999–2004 | appendicular skeletal muscle mass index (ASMI) | Cox proportional hazard regression analysis: HR 0.82 per 1 $kg/m^{2,}$ 95%CI:0.73–0.92 |
| muscle strength | (Jochem et al. 2019) | Meta-Analysis<br>N=39,852 | knee extension strength, knee flexion strength, hand grip strength, | Random-effects meta-regression: Lowest vs. highest category: HR1.80, 95%C: 1.54–2.10 |

| | | | | |
|---|---|---|---|---|
| | | | quadriceps isometric strength, quadriceps maximal voluntary contraction force, Bench press leg press, Medical Research Council scale for muscle strength | |
| hand grip strength | (García-Hermoso et al. 2018) | Meta-Analysis:<br>N=1,907,580<br>N= 63,087 | Hand Grip Strength | Cox proportional hazard regression: HR=0.69; 95% CI, 0.64-0.74 |
| | (Chua et al. 2020) | N=13,789<br>N= 533 | Hand Grip Strength | Cox proportional hazard regression: lowest vs. highest quartile: HR: 2.05, 95%CI:1.44–2.90 |
| timed “Up & Go” | (Ascencio et al. 2022) | N=427<br>N=81 | Timed “Up & Go” | Cox proportional hazard regression: HR = 1.05; 95% CI: 1.02–1.09 |
| | (Chua et al. 2020) | N=13,789<br>N= 533 | Timed “Up & Go” | Cox proportional hazard regression: lowest vs. highest quartile: HR:3.08, 95%CI: 2.17–4.38 |
| gait speed | (Studenski et al. 2011) | Meta-Analysis<br>N=34,485<br>N=17,528 | Gait Speed (distance between 8 ft and 6 m) | HR per 0.1 m/s: 0.88, 95%CI: 0.87-0.90 |
| | (Rolland et al. 2006) | N=7,250<br>N=754<br>older French women enrolled in | 6-m walking speed | Cox proportional hazard regression: highest vs. lowest category: HR: 6.01, 95%CI: 2.81-12.83 |

| | | | | |
|---|---|---|---|---|
| | | Epidémiologie de l'ostéoporose (EPIDOS) | | |
| | (Cooper et al. 2010) | Meta-Analysis<br>N=14 692 | Walking speed | Confounder adjusted, lowest vs. highest quartile: HR 2.87, 95%CI: 2.22 - 3.72 |
| standing balance test | (Cao et al. 2021) | N=5816<br>N=1530 | modified Romberg Test of Standing Balance on Firm and Compliant Support Surfaces | Cox proportional hazard regression: balance impairment vs. no balance impairment HR: 1.44, 95%CI: 1.23-1.69 |
| | (Cesari et al. 2009) | N=3024<br>N=653<br>Health ABC Study | balance (semi- and full-tandem, and single leg stands each held for 30 seconds) tests | Cox proportional hazard regression, confounder adjusted for <53 seconds vs. ≥53 seconds: HR: 1.35, 95%CI: 1.12 - 1.62 |
| frailty index | (Vermeiren et al. 2016) | Meta-Analysis,<br>N=150,763 | 29 different frailty instruments | Meta-Analysis of RR and HR together:<br>Premature mortality: RR 1.83, 95%CI:1.68–1.98 |
| | (Chang and Lin 2015) | Meta-Analysis,<br>N=35,538<br>N=7,994 | Fried's Frailty Phenotype | Cox proportional hazard regression:<br>- robust vs. frail: HR: 2.0; 95% CI: 1.73–2.32<br>- frail vs. pre-frail: HR: 1.48; 95%CI: 1.34–1.63 |
| cognitive health | (Pavlik et al. 2003) | N= 11,444<br>N= 482<br>ARIC Study | DSST | Cox proportional hazard regression: HR per 7-point DSST score increment: 0.86, 95%CI: 0.80-0.93 |
| | (Rosano et al. 2008) | N=3,156<br>N=704<br>Cardiovascular Health Study | DSST | Cox proportional hazard regression: HR: 0.77, 95%CI: 0.68–0.87 |

| | | | | |
|---|---|---|---|---|
| | (Adjoian Mezzaca et al. 2022) | N=5,989<br>National Health and Nutrition Examination Survey | DSST (inverse coded) | Cox proportional hazard regression: HR per 1-SD change in DSST: 1.36, 95%CI: 1.25-1.48 |
| blood pressure | (Satish et al. 2001) | N=12,802<br>EPESE | Systolic Blood Pressure | Cox proportional hazard regression, confounder adjusted:<br>- men 65-84 years old: HR per 10 mmHG increase: 1.04, 95%CI: 1.01-1.07<br>- men >84 years old: HR per 10 mmHG increase: 0.92, 95%CI: 0.86-0.99 |
| | (Todd et al. 2019) | Meta-Analysis<br>N=21,906 | Systolic Blood Pressure | Fixed-effect meta analysis, <140mmHG vs. >140mmHG:<br>- participants with frailty: HR 1.02, 95% CI 0.90 - 1.16<br>- participants without frailty: HR 0.86, 95% CI 0.77 - 0.96 |
| DNA methylation/epigenetic clocks | (Marioni et al. 2015) | Meta-Analysis<br>N=4,658<br>N=862 | $\Delta_{age}$ (Horvath clock and Hannum clock) | Cox proportional hazard regression, confounder adjusted:<br>5-year higher $\Delta_{age}$ (Hannum): HR 1.21, 95%CI: 1.14-1.29<br>5-year higher $\Delta_{age}$ (Horvath): HR 1.09, 95%CI: 1.02-1.15 |
| | (Föhr et al. 2023) | N=395<br>N=187<br>Finnish Twin Study on Aging (FITSA) | GrimAge<br>DunedinPACE | Cox proportional hazard regression, unadjusted:<br>1-SD increase in GrimAge HR: 1.36, 95%CI: 1.18-1.57<br>1-SD increase in DunedinPACE HR: 1.23, 95%CI: 1.05-1.44 |

Supplementary Table 2: Overview of variables available in BASE-II, their respective assessment methods and their inclusion in main and sensitivity analyses.

| Biomarker | BASE-II Variable | Method | Used in |
|---|---|---|---|
| **T0** | | | |
| growth differentiation factor 15 (GDF15) | GDF15 | DNAm-predicted, Illumina MethylationEPIC | Main analysis |
| C-reactive protein (CRP) | CRP | Standard Laboratory, immunoturbidimetry | Main analysis |
| Interleukin-6 (IL 6) | Interleukin-6 (IL 6) | Standard Laboratory, Cytometric Bead Array flex kit | Main analysis |
| Muscle Mass | BMI-standardized appendicular lean mass (ALM) | Hologic® QDR® Discovery™ dual-energy X-ray absorptiometry | Main analysis |
| Muscle Strength | HGS cut-off | Sex- and BMI-specific cut-offs for muscle strength on HGS defined by Fried et al. [1] | Main analysis |
| Hand Grip Strength (HGS) | HGS | Smedley Dynamometer | Main analysis |
| Timed-Up-and-Go (TUG) | Timed-Up-and-Go (TUG) | Clinical Assessment | Main analysis |
| gait speed | 4m gait speed | (time (in s) needed to walk 4 meters) / 4m | Main analysis |
| standing balance test | Tinetti Mobility Test, part 1 | Clinical Assessment | Main analysis |
| frailty index | Fried's Frailty Phenotype | As described in [1] and [2] | Main analysis |
| Cognitive Health | Digit Symbol Substitution Test (DDST) | Neuropsychological Assessment | Main analysis |
| Blood Pressure (BP) | Systolic Blood Pressure (BP) | boso-medicus memory electronic Sphygmomanometer | Main analysis |
| Epigenetic Clock | DunedinPACE | DNAm-predicted, Illumina MethylationEPIC array | Main analysis |
| Epigenetic Clock | DNAmAA Horvath | DNAm-predicted, Illumina MethylationEPIC array | Additional analysis |
| Epigenetic Clock | DNAmAA Hannum | DNAm-predicted, Illumina MethylationEPIC array | Additional analysis |
| Epigenetic Clock | DNAmAA PhenoAge | DNAm-predicted, Illumina MethylationEPIC array | Additional analysis |
| Epigenetic Clock | DNAmAA GrimAge | DNAm-predicted, Illumina MethylationEPIC array | Additional analysis |
| Epigenetic Clock | DNAmAA GrimAge2 | DNAm-predicted, Illumina MethylationEPIC array | Additional analysis |
| Epigenetic Clock | DNAmAA PCHorvath1 | DNAm-predicted, Illumina MethylationEPIC array | Additional analysis |
| Epigenetic Clock | DNAmAA PCHorvath2 | DNAm-predicted, Illumina MethylationEPIC array | Additional analysis |

| | | | |
|---|---|---|---|
| Epigenetic Clock | DNAmAA PCHannum | DNAm-predicted, Illumina MethylationEPIC array | Additional analysis |
| Epigenetic Clock | DNAmAA PCPhenoAge | DNAm-predicted, Illumina MethylationEPIC array | Additional analysis |
| Epigenetic Clock | DNAmAA PCGrimAge | DNAm-predicted, Illumina MethylationEPIC array | Additional analysis |
| Epigenetic Clock | DNAmAA 7-CpG | DNAm-predicted, MS-SNuPE | Additional analysis |
| **T1** | | | |
| Insulin-Like Growth Factor 1 (IGF-1) | Insulin-Like Growth Factor 1 (IGF-1) | Dried Blood Spots (VITAS Analytical Services) | Main analysis |
| Growth Differentiation Factor 15 (GDF 15) | Growth Differentiation Factor 15 (GDF 15) | DNAm-predicted, Illumina MethylationEPIC array | Main analysis |
| C-reactive protein (CRP) | CRP | Standard Laboratory, immunoturbidimetry | Main analysis |
| | hsCRP | Dried Blood Spots (VITAS Analytical Services) | Sensitivity analysis |
| Interleukin-6 (IL 6) | Interleukin-6 (IL 6) | Dried Blood Spots (VITAS Analytical Services) | Main analysis |
| Muscle Mass | BMI-standardized appendicular lean mass (ALM) | Hologic® QDR® Discovery™ dual-energy X-ray absorptiometry | Main analysis |
| Muscle Strength | HGS cut-off | Sex- and BMI-specific cut-offs for muscle strength on HGS defined by Fried et al. [1] | Main analysis |
| Hand Grip Strength (HGS) | HGS | Smedley Dynamometer | Main analysis |
| Timed-Up-and-Go (TUG) | Timed-Up-and-Go (TUG) | Clinical Assessment | Main analysis |
| standing balance test | Tinetti Mobility Test, part 1 | Clinical Assessment | Main analysis |
| frailty index | Fried’s Frailty Phenotype | As described in [1] and [2] | Main analysis |
| Cognitive Health | Digit Symbol Substitution Test (DDST) | Neuropsychological Assessment | Main analysis |
| Blood Pressure (BP) | Systolic Blood Pressure (BP) | boso-medicus memory electronic Sphygmomanometer | Main analysis |
| Epigenetic Clock | DunedinPACE | DNAm-predicted, Illumina MethylationEPIC array | Main analysis |
| Epigenetic Clock | DNAmAA Horvath | DNAm-predicted, Illumina MethylationEPIC array | Sensitivity analysis |
| Epigenetic Clock | DNAmAA Hannum | DNAm-predicted, Illumina MethylationEPIC array | Sensitivity analysis |
| Epigenetic Clock | DNAmAA PhenoAge | DNAm-predicted, Illumina MethylationEPIC array | Sensitivity analysis |
| Epigenetic Clock | DNAmAA GrimAge | DNAm-predicted, Illumina MethylationEPIC array | Sensitivity analysis |

| | | | |
|---|---|---|---|
| Epigenetic Clock | DNAmAA GrimAge2 | DNAm-predicted, Illumina MethylationEPIC array | Sensitivity analysis |
| Epigenetic Clock | DNAmAA PCHorvath1 | DNAm-predicted, Illumina MethylationEPIC array | Sensitivity analysis |
| Epigenetic Clock | DNAmAA PCHorvath2 | DNAm-predicted, Illumina MethylationEPIC array | Sensitivity analysis |
| Epigenetic Clock | DNAmAA PCHannum | DNAm-predicted, Illumina MethylationEPIC array | Sensitivity analysis |
| Epigenetic Clock | DNAmAA PCPhenoAge | DNAm-predicted, Illumina MethylationEPIC array | Sensitivity analysis |
| Epigenetic Clock | DNAmAA PCGrimAge | DNAm-predicted, Illumina MethylationEPIC array | Sensitivity analysis |
| Epigenetic Clock | DNAmAA 7-CpG | DNAm-predicted, MS-SNuPE | Sensitivity analysis |

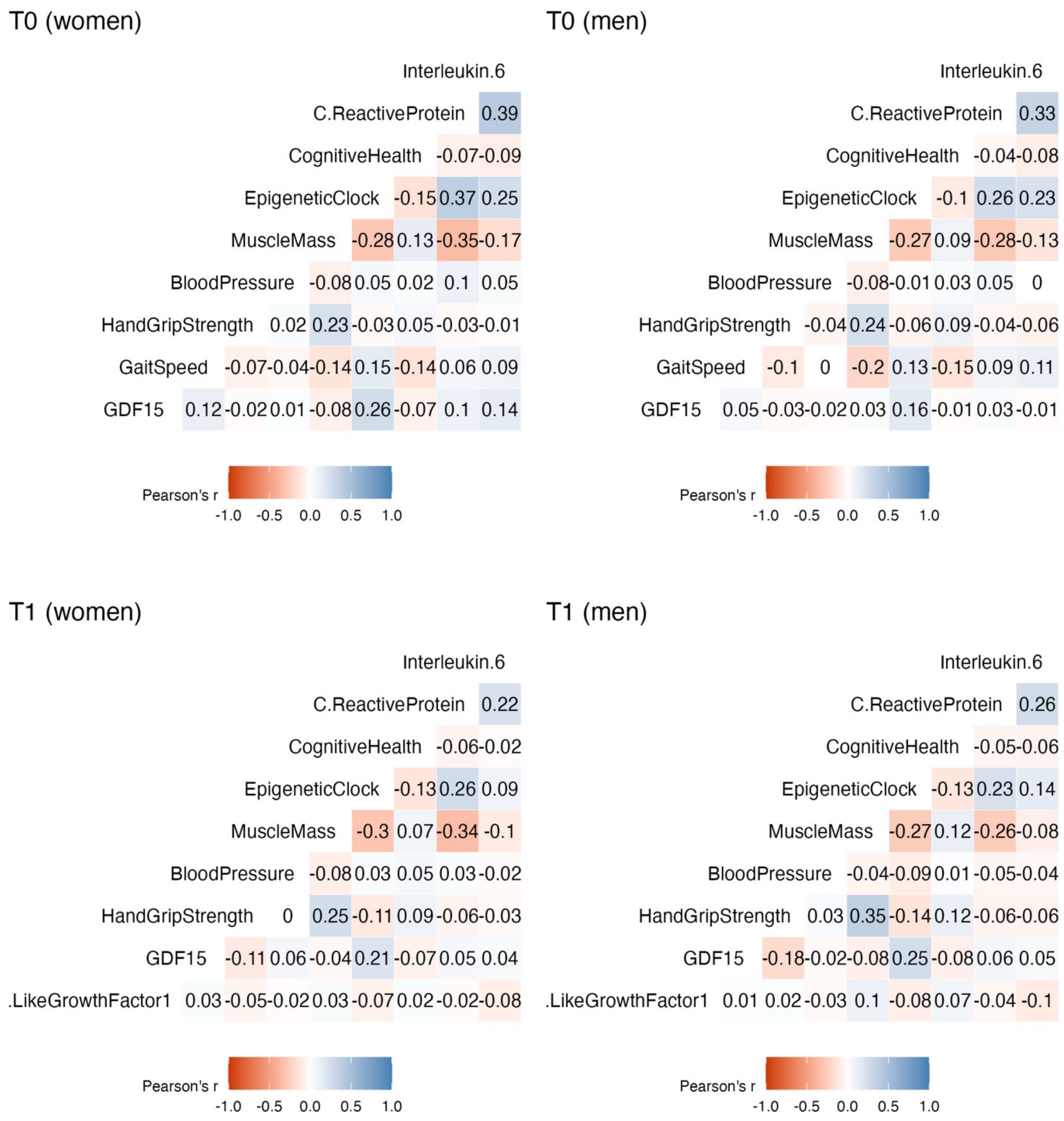


Supplementary Figure 1: Correlation plots (Pearson's r) of continuously scaled biomarkers in sex-stratified subgroups at baseline (T0) and follow-up (T1) using the first imputed dataset.

De Giorgi, A., A. M. Marra, M. Iacoviello, V. Triggiani, G. Rengo, F. Cacciatore, C. Maiello, G. Limongelli, D. Masarone, F. Perticone, P. P. Filardi, S. Paolillo, A. Mancini, M.

Volterrani, O. Vriz, R. Castello, A. Passantino, M. Campo, P. A. Modesti, A. Salzano, R. D'Assante, M. Arcopinto, V. Raparelli, F. Fabbian, A. Sciacqua, A. Colao, T. Suzuki, E. Bossone and A. Cittadini (2022). "Insulin-like growth factor-1 (IGF-1) as predictor of cardiovascular mortality in heart failure patients: data from the T.O.S.CA. registry." Intern Emerg Med **17**(6): 1651-1660 DOI: 10.1007/s11739-022-02980-4.
de Santana, F. M., M. O. Premaor, N. Y. Tanigava and R. M. R. Pereira (2021). "Low muscle mass in older adults and mortality: A systematic review and meta-analysis." Experimental Gerontology **152**: 111461 DOI: https://doi.org/10.1016/j.exger.2021.111461.
Föhr, T., K. Waller, A. Viljanen, T. Rantanen, J. Kaprio, M. Ollikainen and E. Sillanpää (2023). "Mortality Associations With DNA Methylation-Based Biological Aging and Physical Functioning Measures Across a 20-Year Follow-up Period." The Journals of Gerontology: Series A **78**(8): 1489-1496 DOI: 10.1093/gerona/glad026.
García-Hermoso, A., I. Cavero-Redondo, R. Ramírez-Vélez, J. R. Ruiz, F. B. Ortega, D.-C. Lee and V. Martínez-Vizcaíno (2018). "Muscular Strength as a Predictor of All-Cause Mortality in an Apparently Healthy Population: A Systematic Review and Meta-Analysis of Data From Approximately 2 Million Men and Women." Archives of Physical Medicine and Rehabilitation **99**(10): 2100-2113.e2105 DOI: https://doi.org/10.1016/j.apmr.2018.01.008.
Jochem, C., M. Leitzmann, K. Volaklis, D. Aune and B. Strasser (2019). "Association Between Muscular Strength and Mortality in Clinical Populations: A Systematic Review and Meta-Analysis." Journal of the American Medical Directors Association **20**(10): 1213-1223 DOI: https://doi.org/10.1016/j.jamda.2019.05.015.
Lee, J. K., R. Bettencourt, D. Brenner, T.-A. Le, E. Barrett-Connor and R. Loomba (2012). "Association between serum interleukin-6 concentrations and mortality in older adults: the Rancho Bernardo study." PloS one **7**(4): e34218.
Li, Y., X. Zhong, G. Cheng, C. Zhao, L. Zhang, Y. Hong, Q. Wan, R. He and Z. Wang (2017). "Hs-CRP and all-cause, cardiovascular, and cancer mortality risk: A meta-analysis." Atherosclerosis **259**: 75-82 DOI: https://doi.org/10.1016/j.atherosclerosis.2017.02.003.
Marioni, R. E., S. Shah, A. F. McRae, B. H. Chen, E. Colicino, S. E. Harris, J. Gibson, A. K. Henders, P. Redmond, S. R. Cox, A. Pattie, J. Corley, L. Murphy, N. G. Martin, G. W. Montgomery, A. P. Feinberg, M. D. Fallin, M. L. Multhaup, A. E. Jaffe, R. Joehanes, J. Schwartz, A. C. Just, K. L. Lunetta, J. M. Murabito, J. M. Starr, S. Horvath, A. A. Baccarelli, D. Levy, P. M. Visscher, N. R. Wray and I. J. Deary (2015). "DNA methylation age of blood predicts all-cause mortality in later life." Genome Biology **16**(1): 25 DOI: 10.1186/s13059-015-0584-6.
Pavlik, V. N., S. A. de Moraes, M. Szklo, D. S. Knopman, T. H. Mosley Jr. and D. J. Hyman (2003). "Relation between Cognitive Function and Mortality in Middle-aged Adults: The Atherosclerosis Risk in Communities Study." American Journal of Epidemiology **157**(4): 327-334 DOI: 10.1093/aje/kwf209.
Puzianowska-Kuźnicka, M., M. Owczarz, K. Wieczorowska-Tobis, P. Nadrowski, J. Chudek, P. Slusarczyk, A. Skalska, M. Jonas, E. Franek and M. Mossakowska (2016). "Interleukin-6 and C-reactive protein, successful aging, and mortality: the PolSenior study." Immunity & Ageing **13**(1): 21 DOI: 10.1186/s12979-016-0076-x.
Rahmani, J., A. Montesanto, E. Giovannucci, H. Zand, M. Barati, J. J. Kopchick, M. G. Mirisola, V. Lagani, H. Bawadi and R. Vardavas (2022). "Association between IGF-1 levels ranges and all-cause mortality: A meta-analysis." Aging Cell **21**(2): e13540.

Rolland, Y., V. Lauwers-Cances, M. Cesari, B. Vellas, M. Pahor and H. Grandjean (2006). "Physical Performance Measures as Predictors of Mortality in a Cohort of Community-dwelling Older French Women." European Journal of Epidemiology **21**(2): 113-122 DOI: 10.1007/s10654-005-5458-x.
Rosano, C., A. B. Newman, R. Katz, C. H. Hirsch and L. H. Kuller (2008). "Association between lower digit symbol substitution test score and slower gait and greater risk of mortality and of developing incident disability in well-functioning older adults." Journal of the American Geriatrics Society **56**(9): 1618-1625.
Satish, S., D. H. Freeman Jr, L. Ray and J. S. Goodwin (2001). "The relationship between blood pressure and mortality in the oldest old." Journal of the American Geriatrics Society **49**(4): 367-374.
Studenski, S., S. Perera, K. Patel, C. Rosano, K. Faulkner, M. Inzitari, J. Brach, J. Chandler, P. Cawthon, E. B. Connor, M. Nevitt, M. Visser, S. Kritchevsky, S. Badinelli, T. Harris, A. B. Newman, J. Cauley, L. Ferrucci and J. Guralnik (2011). "Gait Speed and Survival in Older Adults." JAMA **305**(1): 50-58 DOI: 10.1001/jama.2010.1923.
Todd, O. M., C. Wilkinson, M. Hale, N. L. Wong, M. Hall, J. P. Sheppard, R. J. McManus, K. Rockwood, J. Young, C. P. Gale and A. Clegg (2019). "Is the association between blood pressure and mortality in older adults different with frailty? A systematic review and meta-analysis." Age and Ageing **48**(5): 627-635 DOI: 10.1093/ageing/afz072.
Vermeiren, S., R. Vella-Azzopardi, D. Beckwée, A.-K. Habbig, A. Scafoglieri, B. Jansen, I. Bautmans, I. Bautmans, D. Verté, I. Beyer, M. Petrovic, L. De Donder, T. Kardol, G. Rossi, P. Clarys, A. Scafoglieri, E. Cattrysse, P. de Hert and B. Jansen (2016). "Frailty and the Prediction of Negative Health Outcomes: A Meta-Analysis." Journal of the American Medical Directors Association **17**(12): 1163.e1161-1163.e1117 DOI: https://doi.org/10.1016/j.jamda.2016.09.010.
